# FROM PRIORITIZATION TO ACCESS: DECISION PATHWAYS FOR ESSENTIAL HEALTH TECHNOLOGIES IN SOUTH AFRICA’S PUBLIC SECTOR

**DOI:** 10.64898/2026.06.29.26356812

**Authors:** Trudy D Leong, Natalie Leon, Andrew G Parrish, Lumbwe Chola, Fundile S Gebremedhin, Tendesayi Kufa, Sarvashni Moodliar, Sumaya Dadan, Thandokazi Mvelashe, Andiswa Nene, Tamara Kredo

## Abstract

**Background:** Timely access to essential health technologies depends on aligning evidence-informed adoption with financing, procurement, delivery, monitoring, and learning. We examined how decision pathways shaped selected health technologies’ progression from prioritisation to implementation in South Africa’s public sector, and their implications for access, equity, sustainability, and learning.

**Methods:** We conducted a qualitative, retrospective, multi-case health policy analysis of twenty essential health technologies purposively selected to capture variation in technology type, disease area, delivery platform, adoption trajectory, and implementation outcome. Document review, process mapping, and key informant interviews were used to reconstruct decision pathways. Analysis was guided by the Policy Cycle Framework, Health Policy Triangle, and Health Technology Assessment (HTA) process domains.

**Results:** Decision pathways varied by technology and delivery platform but followed a common sequence from prioritisation and appraisal to policy endorsement, implementation, and limited reassessment. Medicines and vaccines were generally embedded within established national decision structures. Diagnostics required coordination across laboratory, programme, procurement, and service-delivery systems, while medical-device decisions were more decentralised. Upstream appraisal focused on safety, effectiveness, and public health needs; affordability, infrastructure, equity and sustainability were addressed inconsistently. System learning was evident when routine data, pharmacovigilance, programme review, and guideline revision informed post-adoption adaptation. Weak feedback loops limited reassessment of implementation barriers, equity effects, and sustainability, contributing to delays despite policy endorsement.

**Conclusion:** South Africa has formal structures for evidence-informed technology adoption. HTA would be strengthened by treating appraisal as part of lifecycle governance, with earlier alignment between adoption decisions, financing, procurement, implementation readiness, monitoring, and reassessment.

## Introduction

Timely and equitable access to essential health technologies supports universal health coverage (UHC) (1, 2). However, evidence of clinical benefit, regulatory authorisation, and global normative guidance are insufficient on their own to ensure access. Health systems must translate decisions into financing, procurement, delivery, appropriate use, monitoring, and sustained implementation. In many settings, including low- and middle-income countries (LMICs), evidence-informed policy endorsement may not translate into reliable patient access (3–5). Medicines and vaccines tend to have more established decision and delivery platforms, while diagnostics and medical devices may rely more on fragmented institutional responsibilities or implementation arrangements (6–8).

Health technology assessment (HTA) is increasingly recognized as a mechanism for transparent, evidence-informed priority-setting (1, 2, 8, 9). Contemporary definitions frame HTA as a multidisciplinary, lifecycle-oriented, and system-informed process that assesses the value of health technologies across their lifecycle. In principle, HTA can support decisions that are equitable, efficient, affordable, and responsive to evolving evidence and changing system needs (9, 10). In practice, however, HTA is often applied more narrowly as a technical appraisal with weaker or inconsistent links to downstream decisions on financing, procurement, implementation, and reassessment (11, 12). This gap is particularly relevant in LMICs, where HTA systems are still developing, and institutional connections between appraisal, adoption, and implementation are often incomplete (13, 14).

South Africa provides an informative setting in which to examine these dynamics. Since 1996, public-sector medicine selection has been guided by the Standard Treatment Guidelines and Essential Medicines List (STGs/EML) process, supported by the National Essential Medicines List Committee (NEMLC) and grounded in the National Drug Policy (15). Parallel efforts, including the Essential Laboratory List, have sought to promote rational diagnostic use (16). More recently, National Health Insurance reforms and the 2023 National

Department of Health (NDoH) HTA Methods Guide strengthened policy momentum for more explicit, evidence-informed decision-making (17, 18). However, formal policy architecture does not guarantee implementation. South Africa’s health technology decision-making system spans regulatory approval, guideline and essential-list processes, national programmes, financing, procurement, decentralised service delivery, and monitoring systems. These arrangements create opportunities for evidence-informed prioritisation, but also potential disconnects between appraisal, adoption, and patient access (8, 17, 19–21).

This study examined public-sector decision pathways for selected medicines, vaccines, diagnostics, and medical devices in South Africa. We conducted retrospective multi-case health policy analysis using the Policy Cycle Framework and the Health Policy Triangle (HPT), to explore how decision-making structures operated in practice, and how governance, evidence and implementation domains shaped the progression of essential health technologies from prioritization to routine use. This study uses South Africa as an empirical case to explore the emergence of lifecycle HTA, focusing on the extent to which adoption pathways, implementation readiness, and reassessment processes are institutionalised.

Definitions of key terms used throughout the manuscript are provided in Supplement, File 1, Table 1.

**Table 1.** Case characteristics, adoption trajectories, and implementation constraints.

| Case | Category | Clinical area/<br>Indication | Delivery<br>platform | Dominant decision<br>pathway | Adoption<br>trajectory | Main implementation<br>constraint | Outcome summary |
| --- | --- | --- | --- | --- | --- | --- | --- |
| 3HP (TPT) | Medicine | Tuberculosis | PHC / HIV<br>programmes | Formal policy-led | Delayed adoption | Regulatory, procurement,<br>financing alignment | Policy adopted, but rollout delayed due to<br>regulatory, tender and funding constraints |
| Etonogestrel<br>implant | Medicine | Reproductive health | PHC | Formal policy-led | Iterative/conditional | Acceptability, training,<br>service readiness | Initial scale-up followed by decline and<br>policy/programmatic adjustments |
| DMPA-SC | Medicine | Reproductive health | PHC /<br>community | Formal policy-led | Partial/uneven | Supply chain, delivery<br>model | Policy adopted, but procurement process<br>pending |
| DTG in<br>pregnancy | Medicine | HIV | PHC / HIV<br>programmes | Formal policy-led | Iterative/conditional | Safety concerns, policy<br>revision | Policy was revised multiple times as new<br>evidence emerged |
| TAF | Medicine | HIV | PHC / HIV<br>programmes | Formal policy-led | Delayed adoption | Cost, procurement<br>decisions | Evidence supported use, but adoption was<br>delayed due to contextual considerations,<br>affordability and tendering |
| Fluconazole<br>(CM) | Medicine | HIV/TB<br>(opportunistic<br>infections) | Hospital / PHC | Crisis/donor-mediated | Rapid scale-up | Sustainability, integration<br>into routine systems | Rapid uptake through donor support, later<br>integrated into policy |
| TXA for PPH<br>(PHC) | Medicine | Maternal health/<br>PPH | PHC / referral<br>system | Formal policy-led | Partial/uneven | Training, referral<br>systems, supply | Policy inclusion but incomplete uptake at<br>primary healthcare level |
| Noradrenaline | Medicine | Septic shock/ critical<br>care | Hospital | Implementation-led | Non-adoption/<br>constrained | Infrastructure, workforce<br>capacity, cost | Limited evidence, feasibility constraints |
| Methadone<br>OST/OAT | Medicine | Mental health/<br>substance use<br>disorder | PHC /<br>specialised<br>services | Donor/civil society-<br>mediated | Non-adoption/<br>constrained | Financing, policy<br>prioritisation | Evidence and advocacy present, but<br>constrained service delivery readiness limits<br>national scale-up |
| HPV vaccine | Vaccine | Immunisation/<br>cancer prevention | EPI (school-based) | Formal policy-led | Rapid scale-up | Coverage sustainability | Successfully integrated into national<br>immunisation programme using the<br>integrated school health platform |
| QIV | Vaccine | Immunisation/<br>prevent influenza | EPI / targeted<br>groups | Formal policy-led | Iterative/conditional | Cost-effectiveness,<br>prioritisation | Policy adapted over time with evolving<br>recommendations |
| PCV | Vaccine | Child health/ prevent<br>pneumonia | EPI | Formal policy-led | Rapid scale-up | Procurement, financing | Rapid introduction, but constrained by<br>procurement and cost considerations |
| Apnoea<br>monitors | Device | Neonatal care | Hospital | Implementation-led | Partial/uneven | Procurement,<br>maintenance | Variable uptake dependent on facility<br>capacity |
| CPAP | Device | Neonatal/respiratory<br>care | Hospital | Crisis/implementation-led | Rapid scale-up | Infrastructure, oxygen<br>systems | Rapid adoption driven by service need and<br>clinical demand |
| HHFNO | Device | Respiratory care | Hospital | Implementation-led | Iterative/conditional | Infrastructure | Gradual and uneven uptake with ongoing evaluation |
| CrAg screening | Diagnostic | HIV | NHLS laboratory | Platform-led | Rapid scale-up | Integration into algorithms | Successfully scaled through laboratory platforms |
| Viral load testing | Diagnostic | HIV | NHLS laboratory | Platform-led | Rapid scale-up | System capacity, demand management | Established as routine monitoring with strong scale-up |
| COVID-19 Ag-RDTs | Diagnostic | Infectious disease/ COVID-19 screening | PHC / community | Crisis/donor-mediated | Rapid scale-up | Quality assurance, integration | Rapid deployment followed by challenges in standardisation |
| Dual HIV/syphilis POCT | Diagnostic | Maternal health/ HIV + syphilis screening | PHC | Platform/implementation hybrid | Partial/uneven | Training, QA systems | Policy support but uneven implementation |
| TB-LAM | Diagnostic | Tuberculosis +HIV screening | Hospital / PHC | Platform-led | Delayed adoption | Guideline integration, awareness | Slow uptake despite supportive evidence |

Abbreviations: 3HP=3-month once-weekly isoniazid and rifapentine; AgRDT=antigen rapid diagnostic test; CM=cryptococcal meningitis; COVID-19=coronavirus disease 2019; CPAP=continuous positive airway pressure; CrAg=cryptococcal antigen; DMPA-SC=depot medroxyprogesterone acetate subcutaneous; DTG=dolutegravir; EPI=Expanded Programme on Immunisation; HHFNO=heated humidified high-flow nasal oxygen; HIV=human immunodeficiency virus; HPV=human papillomavirus; NHLS=National Health Laboratory Service; OAT=opioid agonist therapy; OST=opioid substitution therapy; PCV=pneumococcal conjugate vaccine; PHC=primary healthcare; POCT=point-of-care test/testing; PPH=postpartum haemorrhage; QIV=quadrivalent influenza vaccine; TAF=tenofovir alafenamide; TB-LAM=tuberculosis lipoarabinomannan; TPT=tuberculosis preventive therapy; TXA=tranexamic acid.

## Methods

### Study design and analytic orientation

We conducted a qualitative, retrospective, multi-case health policy analysis of the introduction, adaptation, adoption, and implementation of essential health technologies in South Africa’s public sector. A comparative case study design was used to support analytic, rather than statistical generalisation across heterogeneous technologies and decision pathways (22).

Three complementary frameworks guided the analysis: the Policy Cycle Framework, the HTA process, and the HPT. The Policy Cycle Framework provided the primary organizing structure, distinguishing agenda-setting and prioritisation, policy appraisal and formulation, adoption or adaptation, implementation, monitoring and evaluation (23, 24). The HPT informed the interpretation of how actors, context, policy content, and process shaped variation across cases (25). HTA was treated descriptively as a set of evidence-informed functions distributed across the policy cycle, rather than as a single formal appraisal step (21). Refer to Supplement File 1, Figures 1a-1c).

**Figure 1.**
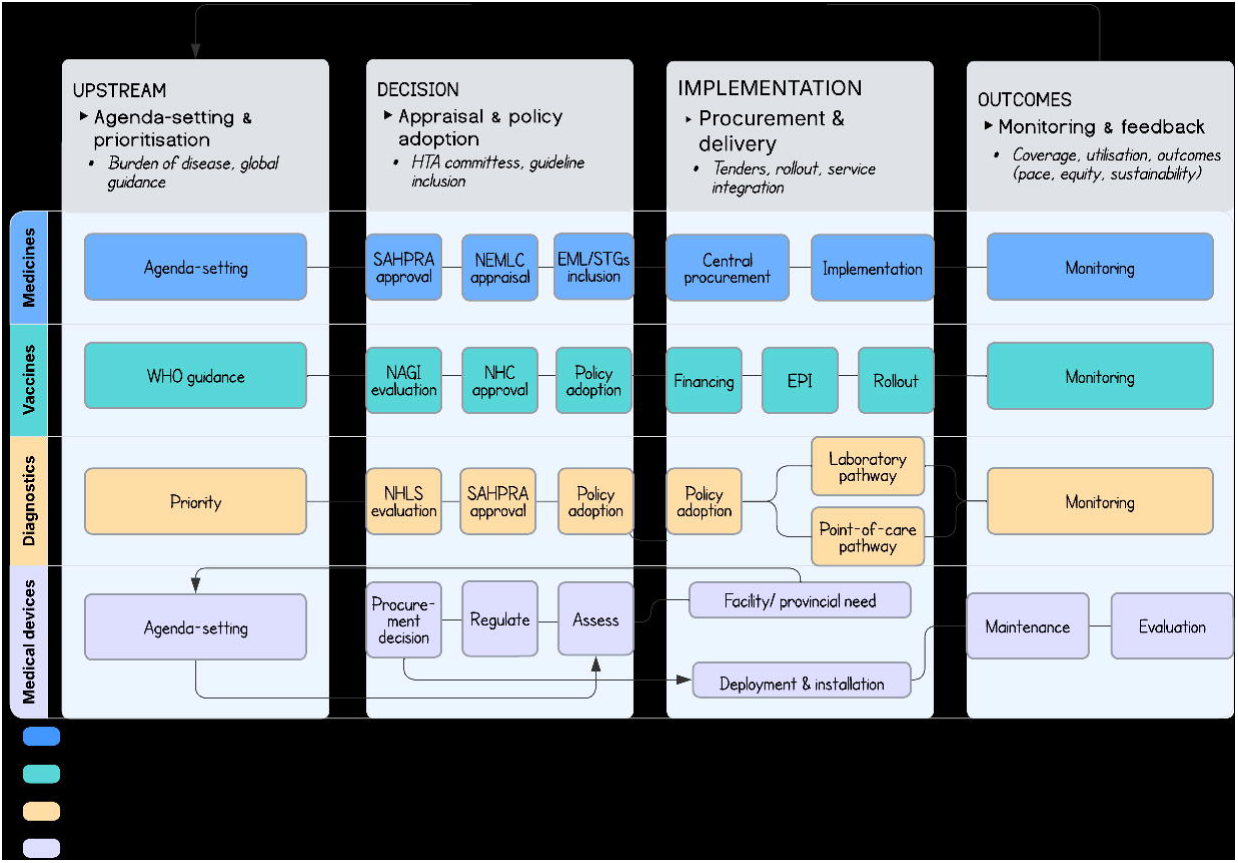
Cross-category framework of health technology adoption pathways in South Africa

### Setting and case selection

The study examined South Africa’s public-sector decision-making processes for primary healthcare (PHC) essential health technologies, focusing on programme delivery, laboratory systems, immunisation, and hospital-based services. We analysed national and subnational institutional arrangements for implementation, including policy development, essential-list processes, regulatory reviews, procurement, clinical training, service readiness, and routine monitoring.

Cases were purposively selected to ensure variation across four technology categories: medicines, vaccines, diagnostics, and medical devices. Selection criteria included disease burden, alignment with national priorities, decision-making salience, delivery platforms, and policy trajectories. Adoption was defined as the process through which technologies were prioritised, assessed, and included or excluded from policy. Implementation referred to integration into routine service delivery, including financing, readiness, procurement, patient access, and monitoring. Medical device cases were identified through consultation with a national policymaker, reflecting the less standardised and inconsistent documentation of these pathways compared with medicines, vaccines, and laboratory diagnostics. The selection aimed to facilitate cross-case comparisons rather than represent all public-sector health technologies.

### Data sources and analysis

Data collection informed process-mapping and health system analysis, focusing on institutional roles, decision interfaces, implementation dependencies, and feedback loops. Documentary sources included national policies, strategy documents, STGs/EML, HTA documents, and implementation circulars. These sources were used to reconstruct policy timelines, identify key decision points, and map institutional roles.

Interviews were conducted between October 2025 and January 2026 with thirteen stakeholders involved in policy development, evidence appraisal, guideline processes, programme management, procurement, laboratory services, and implementation at national, provincial, or district levels. Interviews validated timelines, clarified undocumented processes, and identified implementation constraints. Interviews lasted approximately one hour guided by an interview schedule and summarised using a standardised template (Supplement File 2).

Data extraction used a standardised pre-piloted template covering policy objectives, target populations, agenda-setting drivers, evidence inputs, institutional roles, decision authority, regulatory status, adoption decisions, financing, procurement, implementation requirements, service-readiness constraints, monitoring, timelines, strengths, and proposed system solutions (Supplement File 1, Table 2). Individual case narratives were developed iteratively and refined through triangulation across documentary sources, programme materials, procurement records, and interview accounts. Case summaries are available from Stellenbosch University’s data repository, https://scholardata.sun.ac.za/.

Process maps were developed for each technology category to visualize institutional interfaces, decision points, bottlenecks, delays, and feedback loops (26, 27). Documentary data and interview summaries were analysed using deductive coding informed by the Policy Cycle Framework, HPT, and HTA domains, complemented by inductive coding to identify emergent themes, including political prioritisation, donor influence, crisis-driven adoption, institutional fragmentation, procurement delays, delivery-platform fit, lifecycle support, and variable feedback loops (9, 23, 24, 28).

Analysis proceeded in three stages. First, we reconstructed each technology’s policy and implementation trajectory through within-case analysis. Second, we compared pathways across medicine, vaccine, diagnostic, and medical device categories. Finally, we examined how upstream prioritisation, decision-making coherence, implementation mediators, and downstream consequences influenced adoption, access, equity, sustainability, and learning across technology types. We also assessed whether policy endorsement translated into practical access, and how financing, procurement, service readiness, delivery arrangements, and monitoring influenced this transition.

### Rigour and ethics

Methodological rigour was strengthened through triangulation across documentary and interview data, use of a standardised extraction template, senior authors’ review of extracted data, and maintenance of an auditable link between analytic claims and source material (29). An iterative cross-case analysis examined similarities and differences across technology categories, with emerging interpretations refined by two senior co-authors. Process maps increased transparency by clarifying decision-making and implementation pathways. Divergent cases, including constrained or non-adoption trajectories such as noradrenaline for septic shock and tranexamic acid (TXA) for postpartum haemorrhage (PPH) at primary healthcare (PHC), were analysed to broaden the range of observed outcomes and refine emerging interpretations. Reporting followed established qualitative research standards (29).

The doctoral protocol received approval from Stellenbosch University Health Research Ethics Committee (Ref: S25/03/046). Institutional authorisation was obtained from the National Health Laboratory Service (NHLS) where relevant. All participants provided informed consent. Data were de-identified and stored securely, with access limited to the lead researchers and supervisors.

## Results

We analysed twenty health technology cases across four categories: medicines (n=nine), vaccines (n=three), diagnostics (n=five), and medical devices (n=three). The selected cases spanned HIV, tuberculosis, reproductive health, maternal and child health, immunisation, respiratory care, critical care, and substance-use services, delivered through community-based, PHC, and hospital platforms. Table 1 describes the clinical area, delivery platform, decision pathway, adoption trajectory, implementation constraints, and outcome summary for each case.

Four cross-cutting findings emerged. First, adoption pathways were increasingly formalised, but their institutional location and evidence requirements differed by technology category. Second, evidence was used across cases, although what counted as decision-relevant evidence varied by category and policy stage. Third, adoption translated more plausibly into implementation when appraisal was connected to financing, procurement, service readiness, workforce capacity, quality assurance, and monitoring. Fourth, monitoring and feedback loops were uneven, limiting reassessment and system learning.

## 1. Adoption pathways were formalised unevenly across technology categories

Adoption pathways for technologies showed significant variation across categories. Generally, technologies progressed through a structured policy-adoption continuum: agenda-setting, appraisal, policy formulation, adoption, implementation planning, monitoring, and adaptation. However, the institutional location, sequencing, and accountability for these functions varied substantially by technology category (Supplement File 1, Figures 2–5). Figure 1 integrates these pathways into a cross-category framework, highlighting a shared sequence but divergent arrangements for decision authority, evidence appraisal, implementation responsibility, and feedback.

Medicines had the most established adoption pathway, anchored by regulatory approval from the South African Health Products Regulatory Authority (SAHPRA) and a structured process that included NEMLC appraisal, inclusion/exclusion in STGs/EML, procurement and programme guidance. Despite its stability, this pathway was not linear due to factors such as appeals, donor support, affordability issues, delayed registration, tender cycles, stock-out management, and catalytic technical assistance (Supplement File, Figure 2). Informants described NEMLC processes as “highly structured and methodologically rigorous, [although] complex, rigid, and not user-friendly [when programme] timelines were misaligned with procurement cycles”. Cases like TXA for PPH, three-month weekly rifapentine and isoniazid (3HP) for tuberculosis preventive therapy (TPT), and fluconazole for cryptococcal meningitis showed that adoption was shaped by different combinations of urgency, advocacy, financing, regulatory readiness, and affordability (Table 1). For example, the TXA pathway reflected the influence of maternal mortality concerns and provincial advocacy for earlier access; whereas the 3HP pathway showed how regulatory requirements, high prices, and programme-readiness constraints delayed implementation despite donor support and policy interest.

Vaccines followed a similar formal pathway but were more centred on national immunisation programs. The process involved agenda-setting, NAGI technical advice, NDoH leadership discussions, National Health Council (NHC) endorsement, EPI schedule revision, procurement planning, delivery, safety monitoring, coverage tracking and impact evaluation (Supplement File 1, Figure 3). Compared with medicines, vaccine adoption placed greater emphasis on population-level delivery considerations. Key Informant 11 highlighted that, for high-profile vaccines, “political commitments often precede technical review…[with NAGI later providing] justification and advisory input [for policy formulation]”. The human papillomavirus (HPV) vaccine, pneumococcal conjugate vaccine (PCV), and quadrivalent influenza vaccine (QIV) illustrated how political support, ring-fenced funding, EPI optimisation, affordability, procurement timing, target-group prioritisation and seasonal feasibility shaped adoption.

Diagnostics also adhered to the broad policy continuum, with laboratory-based diagnostics benefiting from NHLS systems for performance evaluation, verification, operational feasibility assessment, procurement, quality assurance, routine data capture and reporting, while point-of-care (POC) testing required more extensive coordination, as illustrated by viral-load (VL) monitoring and reflex cryptococcal antigen (CrAg) screening cases. The diagnostic process map distinguishes laboratory-based pathways from decentralised POC pathways (Supplement File 1, Figure 4). Urine TB-LAM, dual HIV-syphilis POC testing, and COVID-19 antigen rapid diagnostic test (AgRDTs) required wider coordination across programmes, laboratories, facilities, procurement systems, quality-assurance mechanisms, and linkage-to-care pathways.

Unlike medicines, vaccines, and laboratory diagnostics, medical device adoption was not governed through a single national advisory pathway. Decision-making was distributed across clinical, managerial, engineering, procurement, finance, and service-delivery actors, with authority varying by facility, province, budget threshold, and service requirements. The mapped process extended from problem identification and regulatory review to assessment of clinical and technical suitability, budget impact, infrastructure readiness, lifecycle costs, maintenance capacity, procurement, implementation, monitoring, and decommissioning (Supplement File 1, Figure 5). Although these processes enabled operational HTA at facility and provincial levels, they were not systematically linked to national priority-setting, essential technology lists, or formal reassessment. Consequently, adoption of technologies such as apnoea monitors, continuous positive airway pressure (CPAP), and heated humidified high-flow nasal oxygen (HHFNO) systems was driven largely by local service needs, infrastructure, maintenance capacity, affordability, and implementation readiness.

The principal distinction across categories was therefore in their differing context, leadership, evidence used, and connections to implementation and feedback. Medicines followed a centralised regulatory and essential-medicines pathway; vaccines have a programme-integrated immunisation approach; diagnostics rely on laboratory and point-of-care pathways; while devices depend on decentralised lifecycle decisions influenced by service needs, local infrastructure, maintenance, and technical capacity (Figure 1). Key informants reported that medicine adoption followed clear, established processes, whereas pathways for other health technologies were less clearly defined and transparent. This indicates that South Africa’s adoption framework varies in strength across categories. Additionally, civil society’s involvement remains limited, highlighting ongoing issues with inclusiveness and accountability in technology pathways.

## 2. Evidence use was structured, but not always sufficiently system-facing

Evidence-informed adoption decisions for technologies varied in scope and application, with safety, efficacy, effectiveness, disease burden, and public health needs being the most consistently considered factors. Evidence on affordability, procurement feasibility, workforce capacity, platform readiness, and sustainability were less consistently considered upstream, despite their importance in translating adoption into access.

For medicines, the STG/EML process structured evidence synthesis, applying the GRADE (Grading of Recommendations, Assessment, Development, and Evaluation) approach, relying on small technical teams and inter-institutional trust. These accounts suggest a relatively mature appraisal pathway, but one vulnerable to limited technical capacity and misalignment with procurement or programme realities. Some medications, like dolutegravir (DTG), were delayed due to safety concerns in pregnancy, while tenofovir alafenamide (TAF) faced affordability issues. Additionally, the use of noradrenaline for septic shock was limited by infrastructure, workforce, and cost factors.

Vaccine decisions focused on disease burden and population- and programme sustainability, with examples such as HPV and PCV vaccines integrating well into the existing EPI platform. QIV progressed more incrementally through assessments of comparative value, affordability, target-group prioritisation, procurement timing, and seasonal feasibility. Nevertheless, participants suggested that vaccine decisions may be more susceptible to political commitments and global policy momentum; Informants emphasized the importance of evidence-based decision-making, along with the need for decisions to consider the context of the full pathway from appraisal to implementation.

Diagnostics required evidence beyond test accuracy, including clinical utility and workflow considerations. While established laboratory systems supported laboratory-based diagnostics, such as VL testing and reflex CrAg screening, decentralised diagnostics, such as urine TB-LAM and dual HIV–syphilis POC testing, facility-level evidence on workflows, training, quality assurance, data systems, accountability, and clinical action. Key

Informant 13 noted that “we are not doing full HTA [for diagnostics], but we are asking the right questions.” Reflex CrAg screening’s effectiveness was limited when diagnostic identification did not align with consistent clinical follow-up and treatment initiation, highlighting that diagnostic value relies on the quality implementation of the full pathway from test selection to patient management.

For medical devices, decisions prioritised feasibility and lifecycle requirements, with examples showing that device decisions had to account for service needs, technical performance, infrastructure, consumables, maintenance, training, user competence, supplier support, replacement planning, and decommissioning. Provincial and facility stakeholders described practical decision criteria for device adoption, depending on facility capacity, engineering support, maintenance, affordability, and safe implementation, as exemplified in the apnoea monitors, CPAP, and HHFNO cases. Key Informant 4 emphasized, “Do not buy equipment because you have money; buy it because the system needs it.” Key Informant 6 similarly warned that relying on individual clinician opinions or supplier influence can break the system.

Evidence appraisal was most effective for implementation when it considered the entire pathway from recommendation to use. For diagnostics and devices, demonstration studies, pilots, and operational data were crucial as they provided insights into feasibility, workflow, quality assurance, user behavior, maintenance, and implementation risks. As Key Informant 1 stated, “demonstration projects are really the best [for implementation]”. These findings support that HTA should examine not just whether technology works, but the conditions under which it can be effectively procured, delivered, utilized, monitored, and reassessed.

## 3. Implementation readiness mediated the translation of adoption into access

Implementation scale-up was more effective when financing, procurement, service delivery, workforce readiness, and monitoring were addressed early. Technologies embedded in established platforms, such as EPI vaccines (e.g., HPV, PCV) and NHLS diagnostics (e.g., VL testing, CrAg screening), had clearer paths to scale-up, whereas newer decentralised technologies depended on local readiness. For example, TB-LAM and dual HIV-syphilis POCT required facility workflows and training, while CPAP needed oxygen infrastructure and trained staff. The etonogestrel implant illustrated how rapid scale-up can falter due to inadequate training and user acceptability.

Predictable financing was decisive for the HPV vaccine, aided by dedicated funding and a school-based approach. In contrast, DMPA-SC faced delays due to misalignment in regulatory processes, tender-timing and programme integration. As Key Informant 13 pointed out, “if you can’t guarantee supply, rather don’t recommend inclusion”. Key Informant 1 similarly noted that “you can have the best and most robust review process, but if you can’t get the drug, then it’s actually a wasteful process”.

Regulatory and procurement sequencing affected access. For instance, although Umbiflow, a portable Doppler device detecting stillbirth risk through umbilical blood-flow assessment, had strong implementation potential, procurement was hindered by unresolved registration issues. COVID-19 highlighted that emergency coordination can expedite technologies, but also that such gains may not be sustainable. Key Informant 7 described poor “sustainability and foresight [when infrastructure was] used and forgotten [rather than maintained for preparedness]”. Effective implementation readiness links policy endorsement to access, making adoption decisions more credible when they outline delivery models, financing, procurement, training, and monitoring mechanisms.

## 4. Monitoring and feedback loops were uneven, limiting lifecycle HTA

Monitoring and feedback mechanisms were present in several cases, but they were not consistently institutionalised across technology categories. Mature programmes had stronger arrangements. Vaccines, including HPV, PCV, and QIV, benefited from coverage monitoring and surveillance. Laboratory diagnostics, including VL testing and CrAg screening, generated test-volume, quality-assurance, and performance data. These systems created opportunities for adaptive policy, including changes to vaccine products, testing algorithms, or implementation guidance as evidence and system conditions evolved.

Monitoring and feedback mechanisms varied across technology categories, with more mature programs demonstrating stronger arrangements. Vaccines, including HPV, PCV, and QIV, benefited from coverage monitoring and surveillance, while laboratory diagnostics, such as VL testing and CrAg screening, generated valuable data for adaptive policy changes. However, monitoring was weaker for technologies delivered outside established programs, such as medical-device systems, which often did not routinely capture essential usage metrics that could inform national reassessment. Donor-supported interventions also experienced monitoring challenges, exemplified by 3HP scale-up issues, where community demand was disrupted by regulatory and supply issues, and DMPA-SC concerns about pharmacovigilance. Even though laboratory monitoring was strong, as with reflex CrAg screening, clinical follow-up remained weaker, with gaps between identifying CrAg-positive patients and initiating treatment. This illustrates a wider problem: monitoring technology use is not equivalent to monitoring whether use improves outcomes.

Feedback loops between implementation and policy review were inconsistent, with few formalised reassessments across the technology lifecycle. The responsibility for acting on implementation data was often unclear, resulting in fragmented learning. Findings indicate that South Africa’s HTA architecture excels in evidence appraisal for medicines but is more variable for vaccines, diagnostics, and devices. The key challenge is to strengthen appraisal methods and institutionalise linkages between appraisal, financing, implementation, and monitoring.

## Discussion

This study explored twenty essential health technologies that moved from prioritisation to implementation in South Africa’s public sector. Adoption pathways were increasingly formalised, but implementation readiness and reassessment were less consistently institutionalised. These findings suggest that implementation readiness, procurement feasibility, platform fit, financing, and post-introduction learning should be assessed earlier, as part of adoption decisions, rather than treated as downstream operational concerns. Evidence from LMICs similarly shows that successful adoption depends on local implementation conditions, not product performance alone. (8, 30, 31).

Across cases, technologies followed similar, recognisable policy functions: agenda-setting, evidence review, deliberation, endorsement, implementation planning, and monitoring. However, these functions were organised differently across technology categories. Medicines were anchored in regulatory approval and NEMLC STG/EML processes. Vaccines were embedded in immunisation governance, schedule planning, procurement, delivery platforms, and surveillance. Diagnostics depended on laboratory or POC platforms, testing algorithms, quality assurance, data systems, and linkage to care. Medical devices followed more decentralised and lifecycle-dependent pathways, requiring attention to specifications, infrastructure, consumables, maintenance, training, replacement, and safe use.

These findings support HTA as lifecycle governance rather than a discrete appraisal step only. In resource-constrained health systems, the value of a health technology is realised when policy endorsement aligns with financing, procurement, delivery capacity, quality assurance, monitoring, reassessment with disinvestment where appropriate, and accountability (9, 32, 33). A rigorous appraisal may have a limited effect if there are procurement issues, untrained users, or a lack of technology maintenance.

The findings are pertinent to countries seeking to integrate HTA within UHC reforms, as resource constraints, institutional capacities, and system incentives determine whether adopted technologies reach routine care (32, 34–36). While global policy is increasingly viewing HTA as a system-wide function linking priority-setting, financing, and service delivery (8, 9, 21, 35–39), much research still focuses on separate policy stages (25, 40, 41).

By applying the Policy Cycle Framework and HPT, this study highlights how various factors affect the translation of policy into implementation and access (23–25). Different technology categories also show varying needs: while medicines benefit from established national pathways, vaccines faced challenges related to political and fiscal considerations, diagnostics required linkage to care, and medical devices relied on decentralised support and maintenance. Thus, a one-size-fits-all HTA pathway may be insufficient; South Africa may require a national framework with specific requirements for different technology categories.

Civil society, community, and patient perspectives were also insufficiently visible. Although clinical and institutional stakeholders were generally represented, the lack of public engagement may reduce legitimacy, transparency and trust, especially regarding decisions on equity and access (42, 43).

### Policy implications

For South Africa, policy adoption should evolve beyond binary inclusion or exclusion by specifying implementation conditions, such as delivery model, responsible institutions, financing routes, procurement pathways, delivery site-readiness criteria, workforce requirements, quality-assurance arrangements, monitoring indicators, and reassessment triggers. Feasibility assessments need to occur earlier in the process. National guidance is essential for medical devices and decentralised diagnostics, guiding specifications, procurement, and lifecycle management. Monitoring should begin at the introduction stage and consider factors such as functionality, equity, linkage to patient care and outcomes, thereby promoting accountability and ongoing sustainability (44). Additionally, incorporating civil society and community perspectives will enhance legitimacy, improving alignment between policy, implementation, access and uptake (43, 45).

### Strengths and limitations

This analysis drew on case studies across technology categories, combining documentary review and stakeholder interviews, and using established policy-analysis frameworks to examine the pathway from prioritisation to implementation. The comparative design allowed identification of mechanisms that may be relevant beyond South Africa, particularly for health systems seeking to institutionalise lifecycle HTA.

There are several limitations. First, the focus was on adoption and implementation processes rather than patient, population, or health-system outcomes. We therefore did not measure realised access, appropriate use, equity, or health impact. Instead, we analysed institutional arrangements and implementation conditions through which access may be enabled or constrained. Second, cases were purposively selected to capture variation across technology categories and policy trajectories, enabling comparison but limiting the depth of analysis for individual technologies. Third, community, patient, and civil society perspectives were underrepresented, revealing a methodological and empirical gap in HTA decision-making. Lastly, medical device and decentralised implementation pathways were less consistently documented than medicine, vaccine, and laboratory diagnostic pathways. Despite interviews providing some clarity, informal and undocumented practices may not have been fully captured.

## Conclusion

South Africa has an increasingly formal and evidence-informed architecture for adopting essential health technologies. Across twenty cases, however, policy endorsement did not automatically translate into routine use or equitable access. Implementation readiness, procurement feasibility, platform fit, monitoring, and reassessment were decisive but unevenly institutionalised.

Operationalising HTA as lifecycle governance could strengthen the translation of evidence-informed decisions into public value. This will require appraisal processes that explicitly integrate recommendations to financing, procurement, service readiness, quality assurance, monitoring, reassessment, and, where appropriate, disinvestment. For South Africa and other countries pursuing UHC, the key question is not only which technologies should be adopted, but under what conditions they can be delivered, sustained, and reassessed equitably over time.

## Supporting information

Supplement File 2, and will be used for the preprint site

Supplement File 2, and will be used for the preprint site

## Acknowledgements

We sincerely thank all key informants who participated in this study. Ethical approval was granted by the Stellenbosch University Health Research Ethics Committee (Ref: S25/03/046), and we received institutional permission to access relevant laboratory service information from the National Health Laboratory Service (Ref: PR2562944). Participation was entirely voluntary, and all contributors provided informed consent. We are grateful for the time, expertise, and thoughtful reflections shared by informants, as well as for their responsiveness to follow-up queries. Their contributions were essential to understanding the policy and health-system processes described in this work.

We also extend our appreciation to Retsedisitsoe Prudence Mazibuko for her technical support; Marc Brandt for his expert technical insights on medical devices; and Shehani Perera for her editorial support, particularly given her expertise in qualitative research.

This manuscript forms part of the doctoral research of Trudy D. Leong, a PhD candidate in Public Health at Stellenbosch University, under the supervision of Andrew G. Parrish, Lumbwe Chola, and Tamara Kredo.

## AI use declaration

During the preparation of this work, the author(s) used ChatGPT (OpenAI) and Grammarly to assist with language refinement and grammar checking. After using this tool, the author(s) reviewed and edited the content as needed and take full responsibility for the content of the published article.

## Data availability

Data supporting the findings of this study are provided in the Supplement and in the accompanying open-access case-study data report, which will be deposited in SUNScholarData, Stellenbosch University’s institutional research data repository. A persistent DOI will be added to the manuscript once the repository record has been published. Raw interview transcripts are not publicly available to protect participant confidentiality.

