## Supplement File 2, and will be used for the preprint site for "FROM PRIORITIZATION TO ACCESS: DECISION PATHWAYS FOR ESSENTIAL HEALTH TECHNOLOGIES IN SOUTH AFRICA’S PUBLIC SECTOR"

### SUPPLEMENT FILE 1

#### Table 1. Definition of key terms used

| Term/acronym | Definition | Reference |
|---|---|---|
| Adoption | The process through which a health technology is prioritised, assessed, and included in or excluded from policy, guidelines, essential lists, or programme recommendations. | 1,2,3 |
| Agenda-setting | The stage at which a health need, disease burden, service gap, or policy priority is recognised and placed on the decision-making agenda. | 4,5 |
| Appraisal | The structured assessment of evidence on a health technology, including clinical benefit, safety, effectiveness, affordability, feasibility, equity, and health-system implications. | 1,6 |
| Clinical utility | The extent to which a technology, especially a diagnostic test, improves clinical decision-making, patient management, or health outcomes. | 7 |
| Decentralised diagnostics | Diagnostic technologies delivered closer to patients or facilities rather than through centralised laboratory systems, often requiring local workflows, training, quality assurance, and data capture. | 8,9 |
| Delivery platform | The service-delivery arrangement through which a health technology reaches users, such as primary healthcare, hospitals, schools, laboratories, community services, or immunisation programmes. | 3,10 |
| Disinvestment | The planned reduction, replacement, or removal of a health technology when evidence, affordability, safety, implementation performance, or system priorities no longer support its continued use. | 11,12 |
| EML | Essential Medicines List; a list of medicines selected for priority use in the public sector based on evidence, public-health need, affordability, and feasibility. | 13,14 |
| EPI | Expanded Programme on Immunisation; the national immunisation platform responsible for vaccine schedule implementation, procurement planning, delivery, coverage monitoring, and safety surveillance. | 15,16 |
| Essential health technologies | Medicines, vaccines, diagnostics, and medical devices considered important for addressing priority health needs and supporting universal health coverage. | 1,13 |
| Evidence-informed decision-making | A decision-making approach that uses the best available evidence alongside contextual factors such as affordability, feasibility, equity, implementation capacity, and public-health priorities. | 17,18 |
| Feedback loops | Mechanisms through which implementation experience, monitoring data, service constraints, or outcome information inform policy review, adaptation, reassessment, or disinvestment. | 11,12 |
| Financing | The allocation or mobilisation of resources required to procure, implement, sustain, monitor, and reassess a health technology. | 13,19 |
| Health Policy Triangle | A policy-analysis framework that examines how actors, context, policy content, and process shape health policy development and implementation. | 4 |
| Health technology | An intervention used to promote health, prevent, diagnose, treat, monitor, or manage disease; in this study, the term includes medicines, vaccines, diagnostics, and medical devices. | 1 |
| Health technology assessment / HTA | A multidisciplinary, evidence-informed process used to assess the value of health technologies across their lifecycle to support transparent, equitable, efficient, and affordable decision-making. | 1 |
| Implementation | The integration of an adopted health technology into routine service delivery, including financing, procurement, infrastructure, workforce readiness, training, quality assurance, monitoring, and patient access. | 3,10 |
| Implementation readiness | The extent to which the health system is prepared to introduce and sustain a technology, including financing, procurement, workforce capacity, infrastructure, training, quality assurance, and monitoring arrangements. | 10,20 |

| | | |
|---|---|---|
| Institutionalisation | The embedding of decision-making processes, responsibilities, evidence requirements, accountability mechanisms, and feedback loops into routine governance structures. | 4,10 |
| Laboratory-based diagnostics | Diagnostic technologies delivered through established laboratory systems, usually supported by laboratory infrastructure, verification, procurement systems, quality assurance, information systems, and reporting. | 8,21 |
| Lifecycle governance | A governance approach that considers a health technology from prioritisation and appraisal through adoption, procurement, implementation, monitoring, reassessment, and possible disinvestment. | 1,22,23 |
| Lifecycle HTA | The application of HTA functions across the full technology lifecycle rather than as a single technical appraisal event before adoption. | 1,22,23 |
| LMICs | Low- and middle-income countries; countries facing resource constraints and health-system capacity challenges that may affect adoption, implementation, and sustained access to health technologies. | 13,19 |
| Medical devices | Health technologies such as equipment, instruments, or systems used in care delivery, which often require infrastructure, consumables, technical support, maintenance, training, replacement planning, and safe-use systems. | 6,9,22 |
| Monitoring and evaluation | The routine collection and assessment of data on availability, coverage, utilisation, quality, functionality, safety, outcomes, equity, and sustainability after a technology is introduced. | 3,11,12 |
| NAGI | National Advisory Group on Immunisation: a technical advisory body that provides evidence-informed recommendations on vaccine policy and immunisation decisions. | 15,24 |
| NDoH | National Department of Health: the national authority responsible for health policy, programme leadership, guidance, and stewardship in South Africa. | 25 |
| NEMLC | National Essential Medicines List Committee; the committee responsible for reviewing evidence and making recommendations on medicines for inclusion in the Standard Treatment Guidelines and Essential Medicines List. | 14 |
| NHLS | National Health Laboratory Service, the public-sector laboratory service that supports laboratory-based diagnostic testing, verification, procurement, quality assurance, data systems, and reporting. | 21 |
| NHI | National Health Insurance: South Africa's health financing reform intended to support universal health coverage and more equitable access to health services. | 25,26,27 |
| Policy adoption continuum | The sequence of policy functions through which technologies move from agenda-setting and appraisal to policy formulation, adoption or adaptation, implementation planning, monitoring, and reassessment. | 4,5,12 |
| Policy Cycle Framework | A policy-analysis framework that organises decision-making into stages such as agenda-setting, policy formulation, adoption, implementation, monitoring, evaluation, and adaptation. | 4,5 |
| Point-of-care tests / POC diagnostics | Diagnostic tests performed near the patient or at the facility level that require attention to workflows, staff training, quality assurance, data capture, supply chains, and linkage to care. | 8 |
| Procurement | The process of purchasing or securing health technologies and related supplies, including tendering, supplier selection, contracting, distribution, and stock management. | 28,29,30 |
| Quality assurance | Systems and processes that are used to ensure that a health technology is delivered, maintained, interpreted, and used safely, accurately, and consistently. | 8,31 |
| Reassessment | The periodic review of an adopted technology using new evidence, implementation data, affordability considerations, safety information, and health-system performance. | 11,12 |
| Regulatory authorisation | Approval, registration, or exceptional access granted by the relevant regulatory authority before a health technology can be used or procured. | 6,32 |
| SAHPRA | South African Health Products Regulatory Authority, the national regulator responsible for medicines, vaccines, medical devices, and related health products. | 32,33 |
| Section 21 access | A mechanism allowing exceptional access to unregistered health products under defined regulatory conditions in South Africa. | 32 |

| | | |
|---|---|---|
| Service readiness | The availability of the infrastructure, staff, equipment, training, supplies, systems, and operational conditions needed to deliver a health technology safely and effectively. | 10,20 |
| STGs | Standard Treatment Guidelines: clinical guidance used in South Africa's public sector to support evidence-informed, standardised care. | 14,34 |
| STGs/EML process | The combined Standard Treatment Guidelines and Essential Medicines List process used to guide public-sector medicine selection and use. | 14,34 |
| Total Cost of Ownership / TCO | The full direct and indirect costs of a medical device across its lifecycle, including acquisition, implementation, operation, maintenance, monitoring, replacement, and decommissioning. In medical device HTA, TCO should be assessed before adoption because the purchase price alone may underestimate the resources required for safe, equitable, and sustainable implementation. | 29,30,35 |
| Universal health coverage / UHC | The goal that all people should have access to needed health services and technologies of sufficient quality without financial hardship. | 13,19 |

### Table 2. Standardised data-collection template for document review

**INNOVATION:**

| Policy Cycle Stage | Sub-Code | Innovation Type | Extract (Raw Data) | Interpretation / Analytical Memo | Actor(s) Involved | Supporting Policy / Document |
|---|---|---|---|---|---|---|
| **Agenda Setting** | Problem identification | | | | | |
| | Political prioritisation | | | | | |
| | Stakeholder advocacy | | | | | |
| | Collaboration | | | | | |
| **Policy Analysis** | Evidence review | | | | | |
| | Equity & access | | | | | |
| | Feasibility & capacity | | | | | |
| | Acceptability | | | | | |
| | Financial analysis | | | | | |
| | Collaboration | | | | | |
| **Policy Adoption** | Formal approvals | | | | | |
| | Integration into policy instruments | | | | | |
| | Legal/legislative actions | | | | | |
| | Collaboration | | | | | |
| **Implementation** | Procurement & supply chain | | | | | |
| | Service delivery | | | | | |
| | Partnerships | | | | | |
| | Barriers | | | | | |
| **Monitoring and evaluation** | Routine monitoring | | | | | |
| | Performance audits | | | | | |
| | Impact evaluation | | | | | |
| | Feedback loops | | | | | |

**References:**

**Figure 1a. The Policy Cycle Framework**

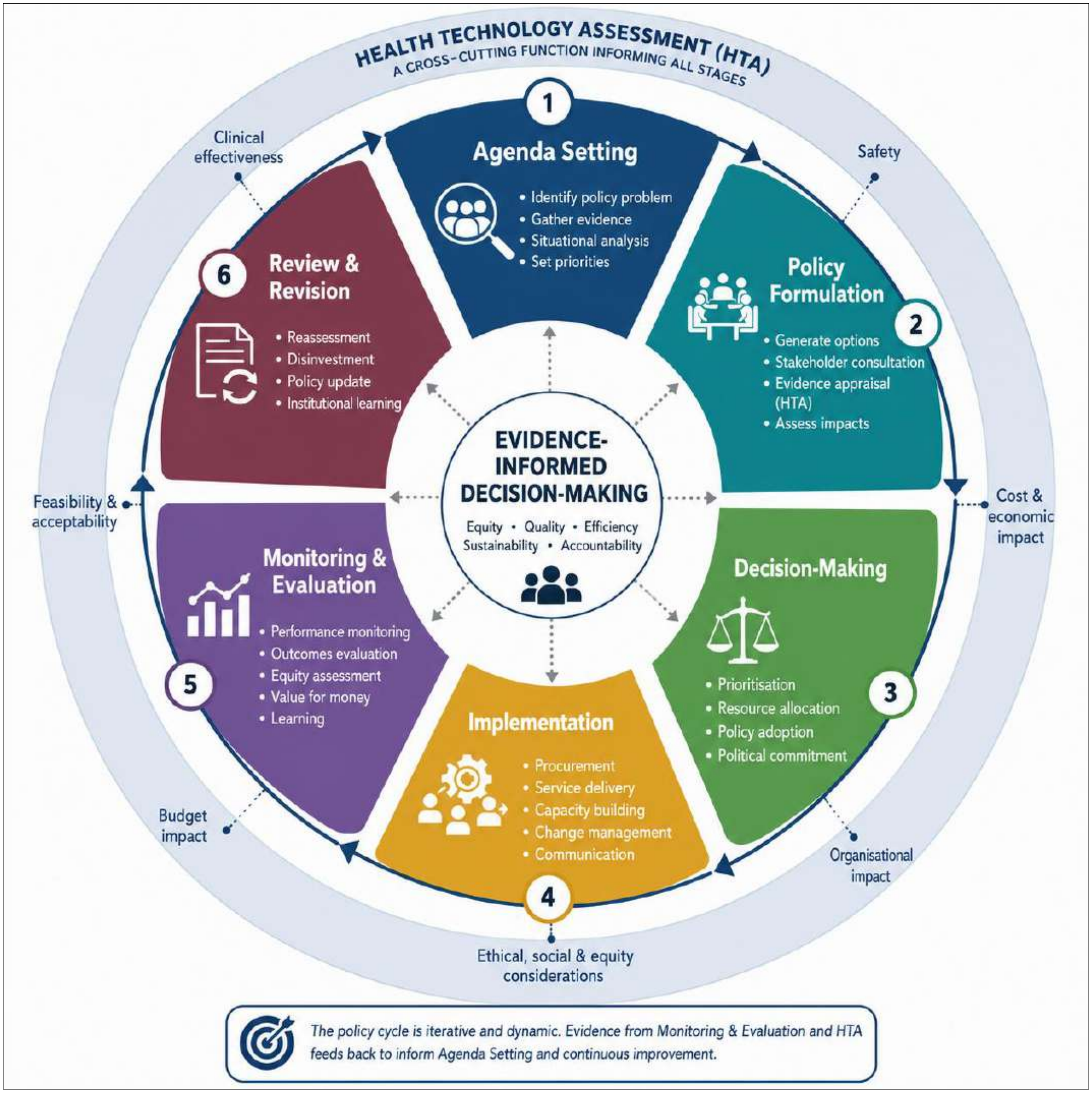

**Figure 1b. The Health Policy Triangle**

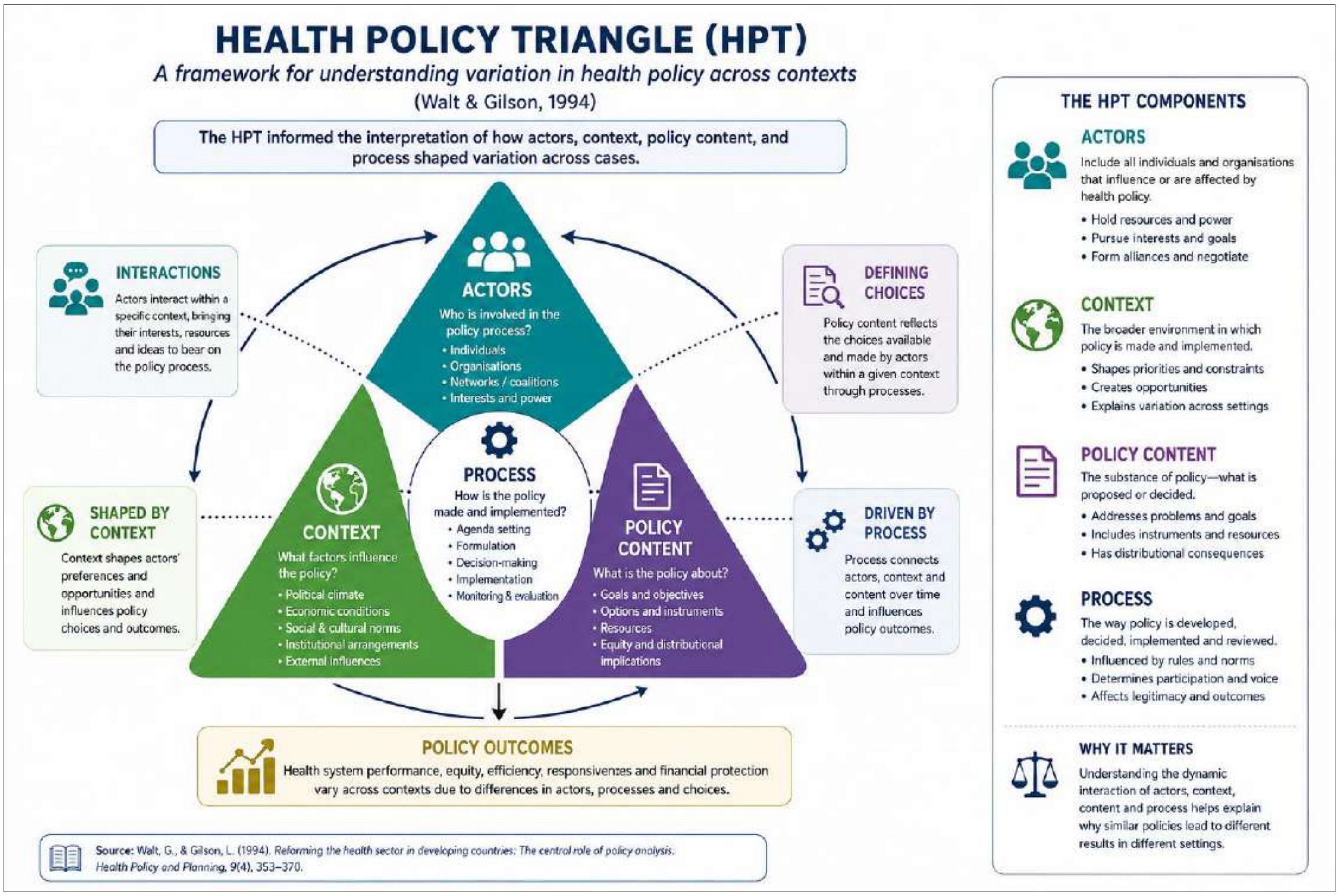


.

**Figure 1c. The Health Technology Assessment Process**

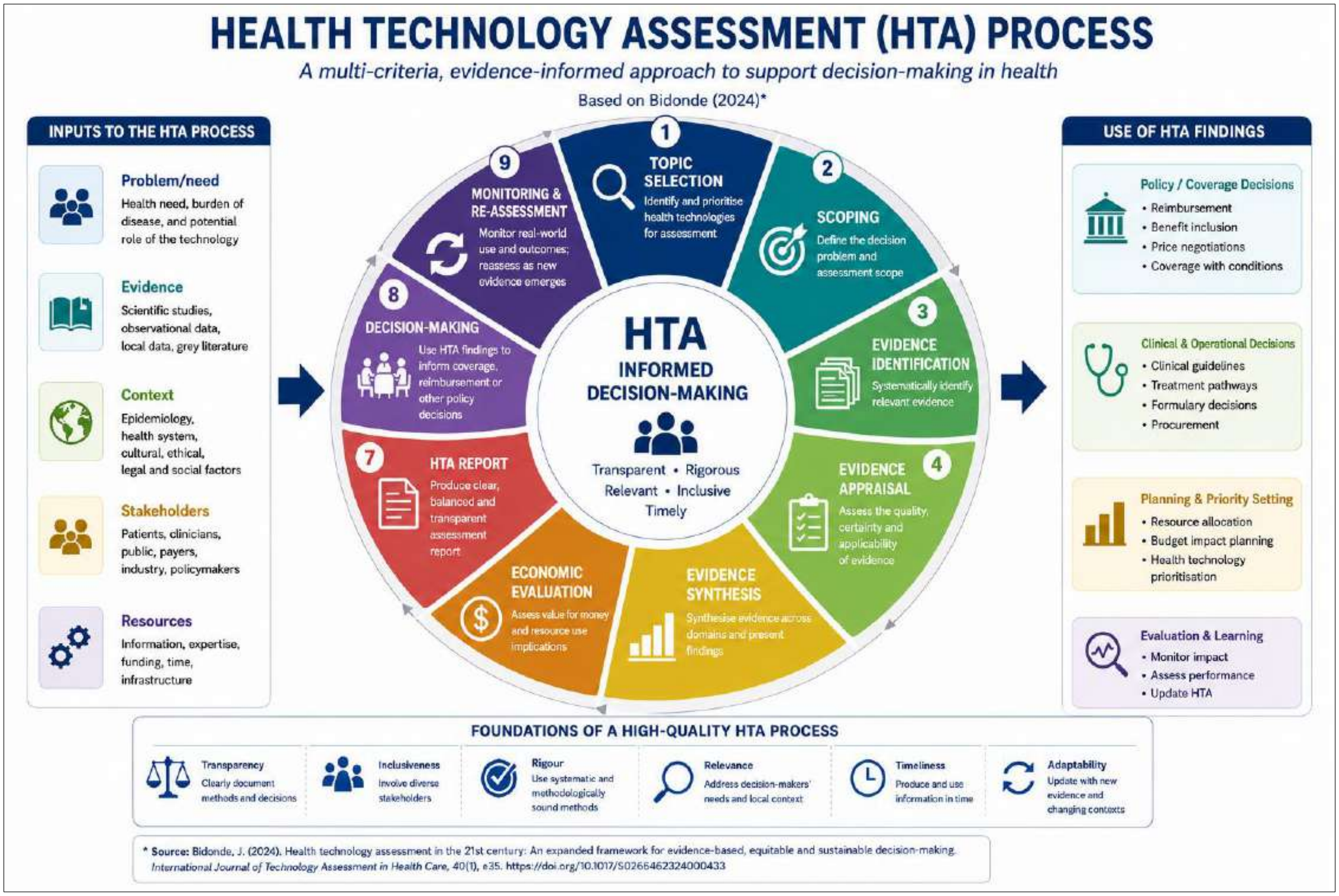


**AI-assisted graphics statement**

Supplementary Figures 1a, 1b, and 1c were created with assistance from ChatGPT for visual layout and presentation only. All scientific content, labels, and data were reviewed, verified, and approved by the authors, who take full responsibility for the final materials.

**Figure 2. Process map for the introduction of medicines in the South African public sector**

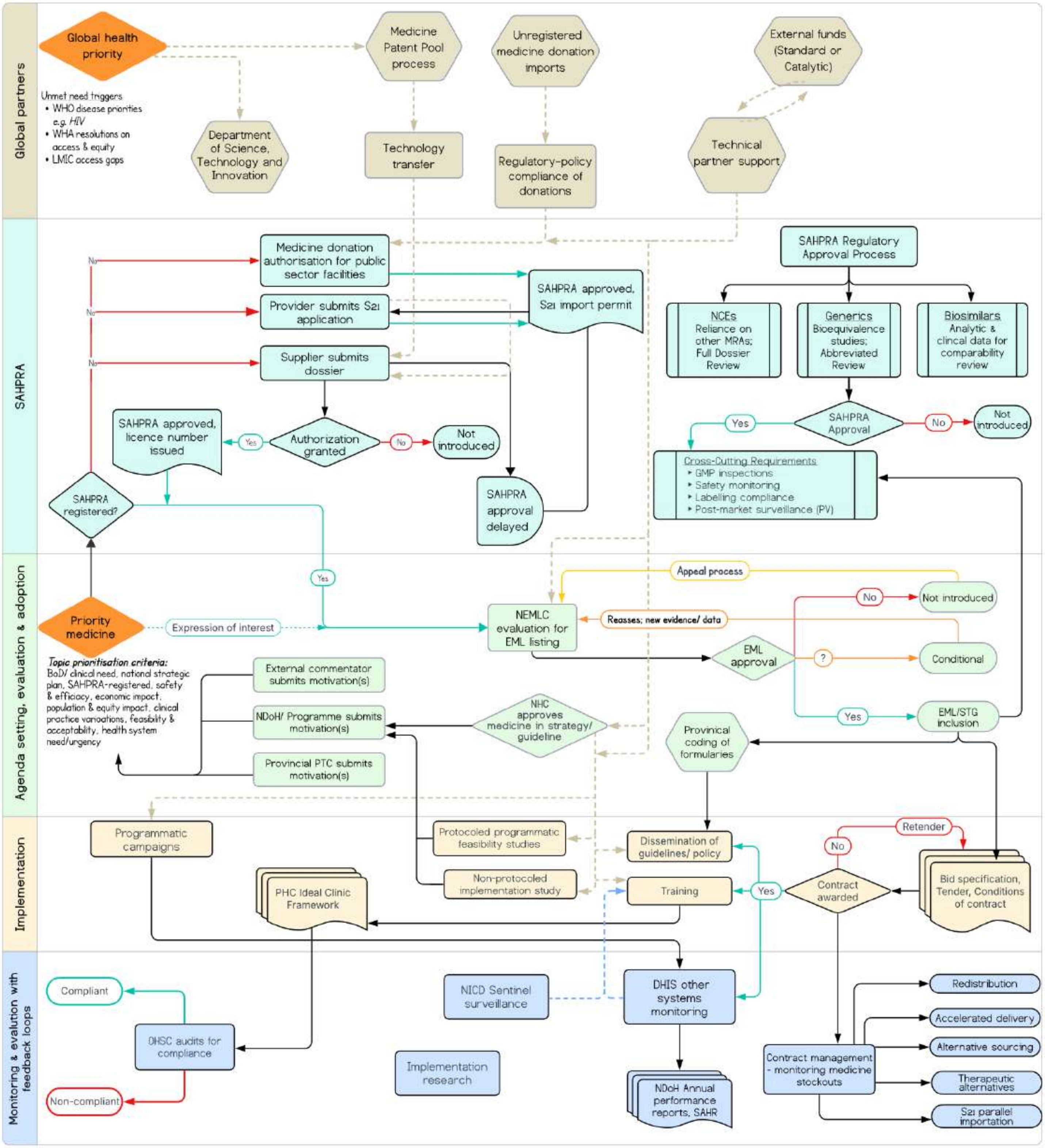


*Abbreviations: BoD=burden of disease; DHIS=District Health Information System; EML/STGs=Essential Medicine List and Standard Treatment Guidelines; NEMLC=National Essential Medicines List Committee; LMIC=Low- and Middle-Income Country; NCD=new chemical entity; NHC=National Health Council; NICD=National Institute for Communicable Diseases; OHSC=Office of Health Standards and Compliance; PHC=Primary Healthcare; PTC=Pharmacy and Therapeutics Committee; S21=Section 21 of the Medicines and Related Substances Act (Act 101 of 1965); SAHPRA=South African Health Products Regulatory Authority; SAHR=South African Health Review; WHA=World Health Assembly; WHO=World Health Organization*

- Medicines generally followed formal regulatory, guideline/EML, procurement, and programme implementation processes, although adoption did not consistently translate into timely or complete scale-up.

**Figure 3. Process map for the introduction of vaccines in the South African public sector**

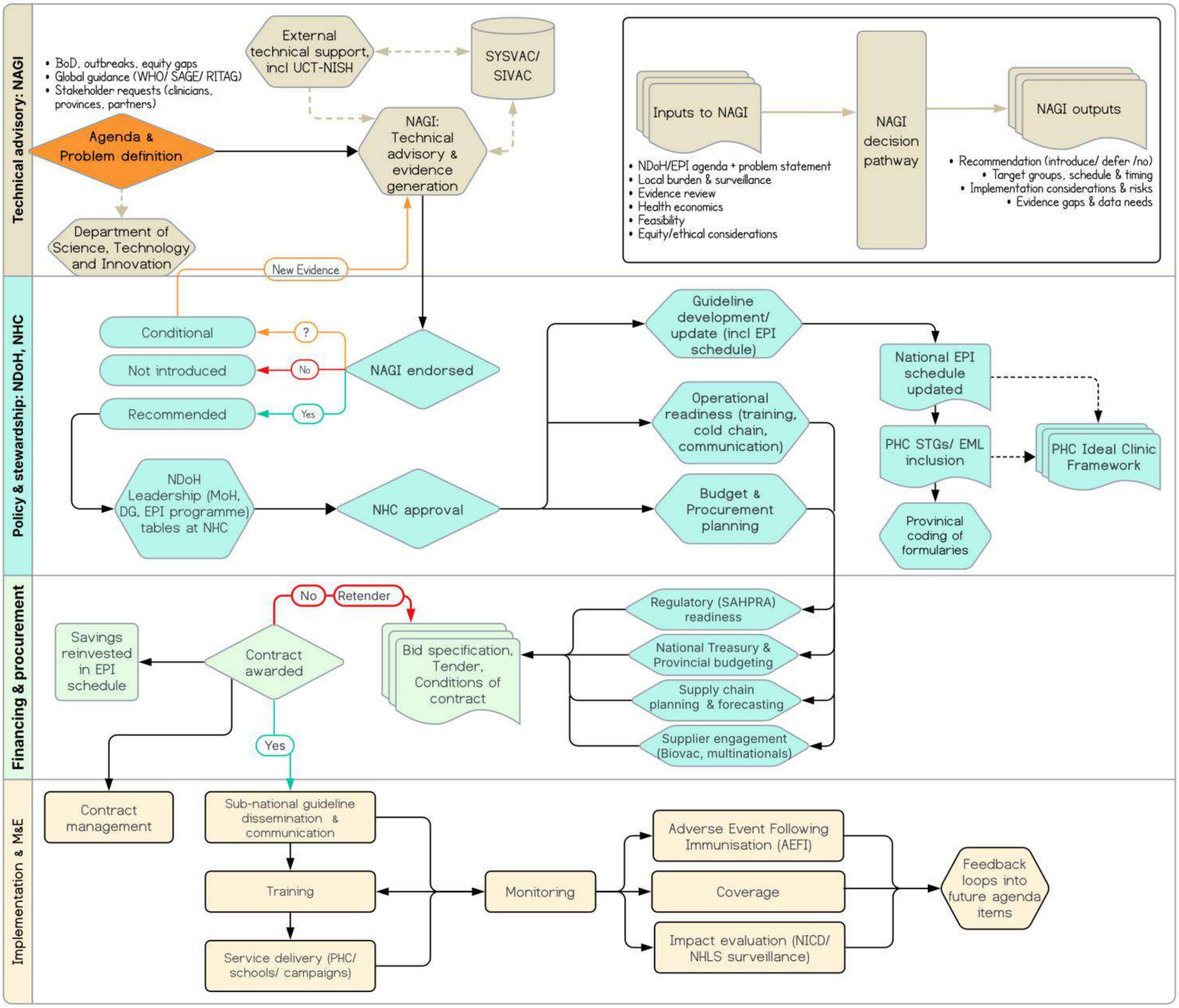


*Abbreviations: AEFI=Adverse Event Following Immunisation; BoD=burden of disease; DG=Director General; EPI=Expanded Programme on Immunisation; MoH=Minister of Health; NAGI= National Advisory Group on Immunisation; NHC=National Health Council; NHLS=National Laboratory Services; NICD=National Institute for Communicable Diseases; PHC=Primary Healthcare; RITAG=Regional Immunisation Technical Advisory Group, SAGE=Strategic Advisory Group of Experts on Immunization to the WHO; SAHPRA=South African Health Products Regulatory Authority; STGs/EML= Standard Treatment Guidelines and Essential Medicines List; SIVAC=Supporting Independent Immunization and Vaccine Advisory Committees Initiative; SYSVAC=Systematic Reviews of Vaccines (project by WHO, to support NITAGs including NAGI with technical evidence); UCT-NISH= University of Cape Town National Immunisation Safety Hub (AEFI monitoring, vaccine economics & policy training); WHO=World Health Organization*

- Vaccines followed a structured immunisation pathway linking technical appraisal, financing, procurement, and delivery through the Expanded Programme on Immunisation.

**Figure 4. Process map for the introduction of diagnostics in the South African public sector**

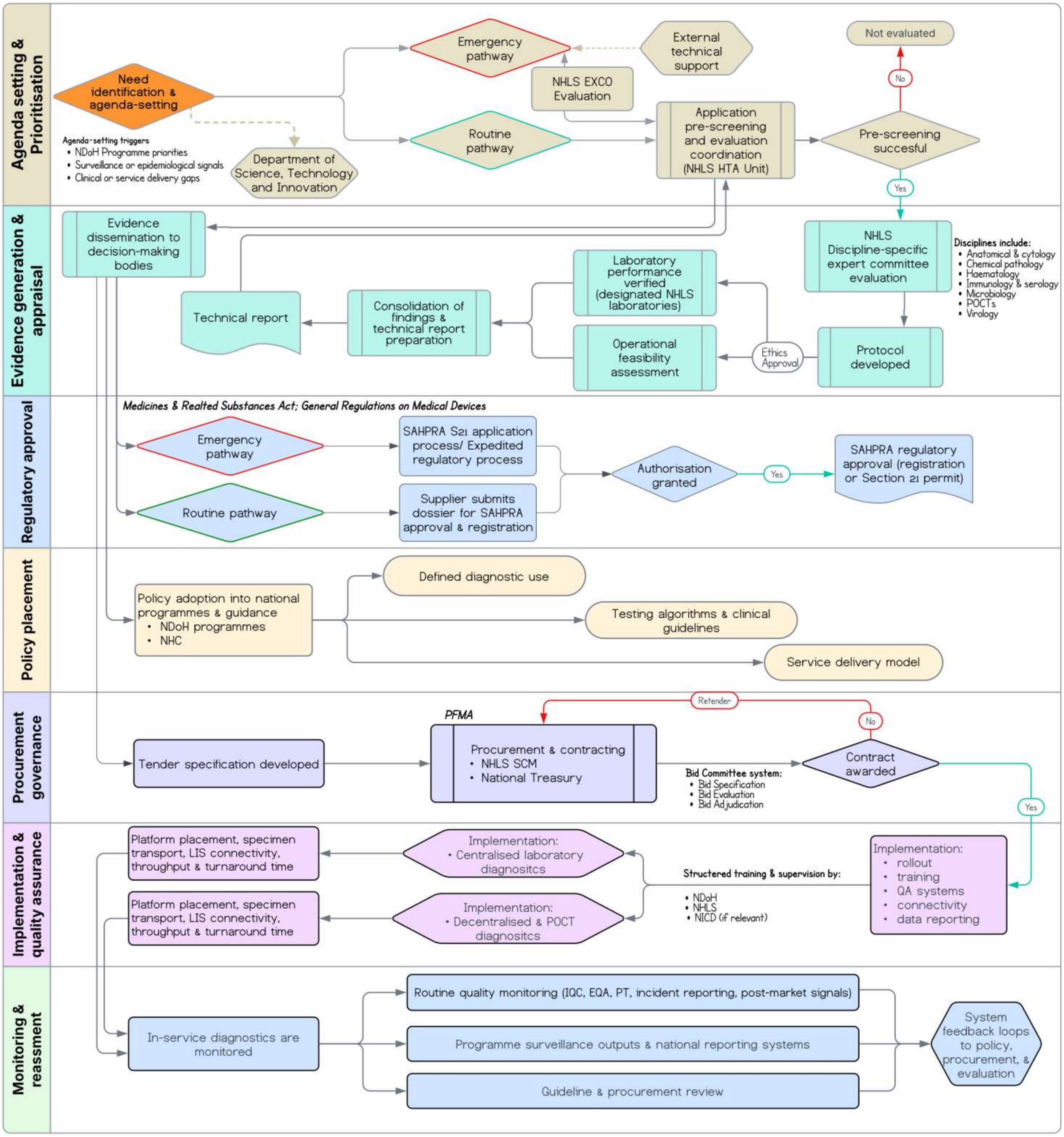


*Abbreviations: EQA=external quality assessment; EXCO=Executive Committee; IQC=internal quality control; LIS=laboratory information system; NDoH=National Department of Health; NHC=National Health Council; NHLS=National Health Laboratory Service; NICD= National Institute for Communicable Diseases; PFMA=Public Finance Management Act; POCT=point-of-care testing; PT=proficiency testing; QA=quality assurance; SAHPRA=South African Health Products Regulatory Authority; SCM=supply chain management*

- Diagnostics followed platform-dependent pathways shaped by guideline processes, laboratory or point-of-care systems, procurement, quality assurance, training, and data integration.

**Figure 5. Process map for the introduction of medical devices in the South African public sector**

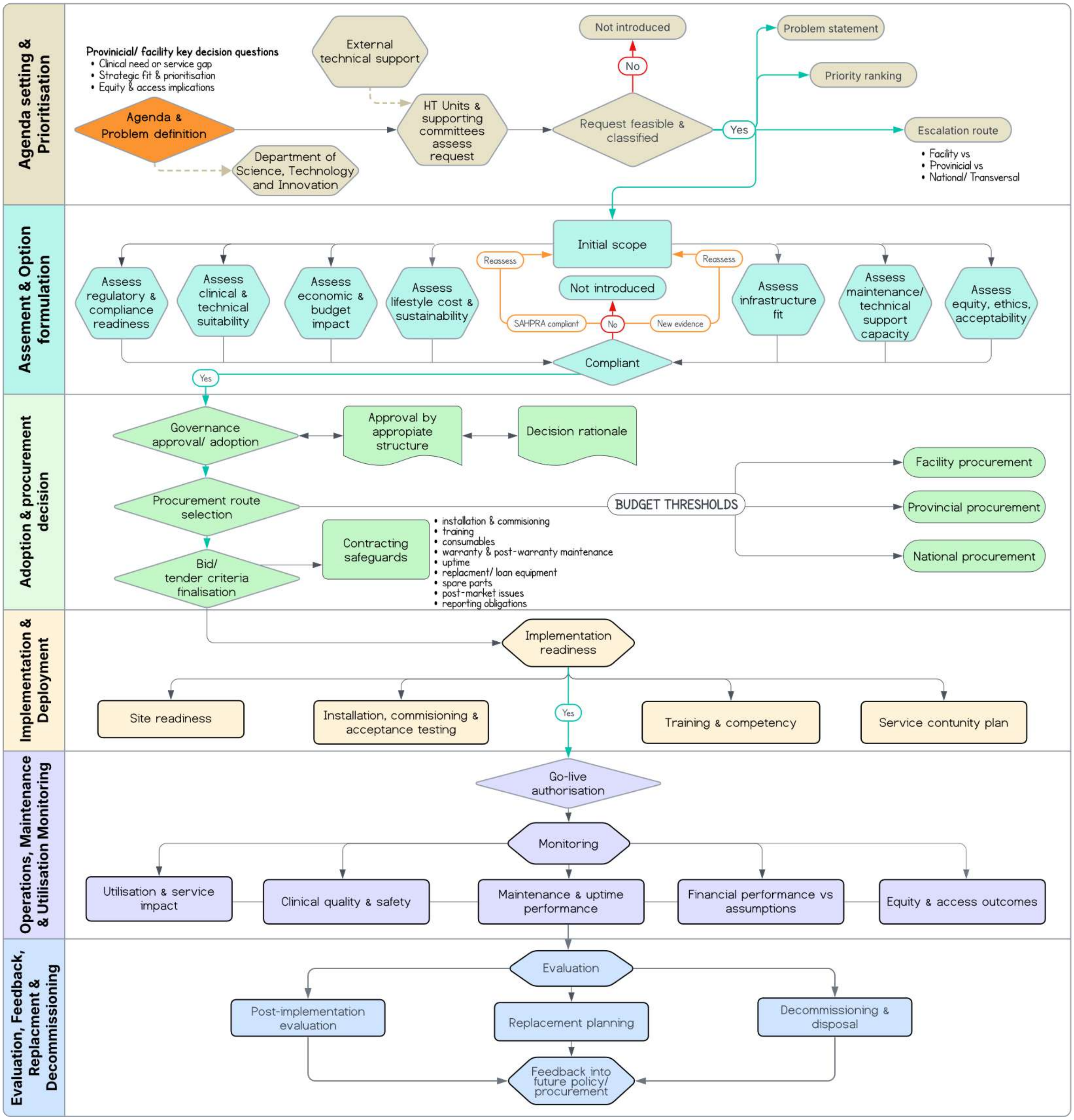


*Abbreviations: SAHPRA=South African Health Products Regulatory Authority*

- Medical devices followed more decentralised, implementation-led pathways dependent on service demand, infrastructure, procurement, maintenance, workforce capacity, and facility readiness.
