## Supplement File 2, and will be used for the preprint site for "FROM PRIORITIZATION TO ACCESS: DECISION PATHWAYS FOR ESSENTIAL HEALTH TECHNOLOGIES IN SOUTH AFRICA’S PUBLIC SECTOR"

### A: INTERVIEW SCHEDULE

Welcome the participant to the meeting. Introduce yourself and anyone with you from the research team. Clarify that the information provided is clear and that consent has been signed. Request permission to start recording. Reassure that confidentiality is adhered to and that we can discuss if there are any concerns at any time during the interview. The interview should take 45 to 75 minutes.

Introduce the concept of the Health Technology Assessment (HTA) process, decision-making for topic prioritisation and selection of essential medicines, devices and diagnostics for primary care settings in the South African public sector.

Below are some questions that we will reflect on for improved decision-making for the selection of essential medicines, devices or diagnostics in primary care settings.

#### Demographic Information

| |
|---|
| Work Location |
| Describe your role in the health sector? |
| How long have you worked in this field? Any other relevant work experience you wish to share? |

#### Draft schedule of questions

An interview guide with seven broad guiding questions will be used, appropriately selected for each stakeholder group (including senior and middle-level policymakers from the public sector, professional association representatives active in health policy reform, health system researchers, from academia and other research organisations, non-state sector representatives active in health policy reform, PHC service providers, and representatives of advocacy groups).

(1) *Process:* As one of the participants contributing directly (through NEMLC membership)/ indirectly (through peer review) to the decisions for selecting essential medicines, devices and diagnostics for PHC (during the PHC STGs and EML review period of 2020 to 2024), briefly describe what this entailed.

(2) *NDoH guides/policies:* Are you aware of the piloted topic prioritisation framework for HTA evaluations and the NDoH HTA Methods Guide? What is your understanding and opinion/perception of these approaches for selecting essential medicines, devices and diagnostics for use at PHC in the public sector?

(3) *Stakeholder engagement:* Are you aware of other participants/groups involved in developing the topic prioritisation and selection of health technology policy/ies, and the decision-making for selecting essential medicines, devices and diagnostics for PHC, and what roles did they play? (Note: Confidentiality of all parties will be maintained).

(4) *Processes and priorities:* How did you and those who participated in or contributed to policy-making for PHC essential medicines, devices and diagnostics go about achieving prioritisation of a topic/s (agenda-setting) and inclusion/deletion or amendment of medicine(s) or diagnostic(s) in the PHC essential lists? Was good governance maintained and how could this be improved to minimise the risk of corruption and improve healthcare access for the more vulnerable populations?

(5) *Implementation of the HTA process:* What facilitators and barriers did you perceive in the review of the 2024 PHC STGs and EML, and what areas do you think should be improved?

(6) *Power dynamics:* Can you describe decision-making dynamics within the NEMLC? Or what is your perception?

- Can you recall and reflect on cases where your or others' voices were not heard, or where groupthink dominated (i.e. others or your decisions "conformed" to pressure from the majority, or "obeyed" the views of a figure perceived to be in a more powerful or authoritative position?
- Can you recall or reflect on decisions where the persuasion from the minority influenced the final decision due to the psychosocial impact of status and power?

(7) *Adoption of recommendation(s):* How was it made clear that a decision had been reached, and were there formal ways of identifying this (e.g. a vote)? Are the final recommendations easily accessible to end-users (including healthcare workers and patients)?

What for you was the most memorable aspect of being part of these decision-making processes? And why?

If you were to write a guide for other decision-makers doing this kind of work- what are one ro tow main "Must do's" one or two " Must not do's" ?

Questions:

Are there any comments you would like to add, including perhaps areas you think we should explore further in these interviews going forward?

Any questions from your side?

*Note:* Document policy analysis will further inform the interview schedule and other potential areas that may need to be probed during the interviews.

Thank you very much for being so responsive and for the generosity of sharing your time.

Note: The interviews were conducted as part of a doctoral study approved by the Stellenbosch University Health Research Ethics Committee (Ref: S25/03/046).

### B: INTERVIEW SUMMARY TEMPLATE

- For completion within 36 hours of conducting the interview, 2+ page summary of key ideas emerging from the interview in relation to your research question
- Write the notes based on your written interview notes and your memory
- This summary becomes your first level of analysis of interview content

INTERVIEW ID:

| Respondent: # | Interviewer: |
| --- | --- |
| Date of interview: | Research context: |
| Location: Zoom | Summary by: X (name) on Y (date) |

Demographics

- Gender:
- Professional background:

Main issues raised in the interview

- Role in the policy decision-making of HTA innovation introduction, integration & implementation *(Agenda setting, Evidence review and policy formulation, Implementation of policy, Monitoring & evaluation)*
- Reflections on the experience of participating in the policy decision-making of HTA innovation
- Facilitators and barriers identified in the policy-making experience
- Specific insights re Lessons learnt
- Recommendations made for improving future policy decision-making pathways

Main gaps to explore further in other interviews

Memorable quotes

- In written notes, indicate a time stamp on the digital recording to guide where to find potentially good quotes

Questions/comments from participant:

My reflections

- What stood out for me about the content shared?
- What worked well, less well in the interview (incl. my questions/reflections/feelings about it, what to learn from my experience for the next interview)
